# Prevalence of influenza vaccination among adults with high-risk conditions, United States, 2019

**DOI:** 10.1101/2020.12.21.20248635

**Authors:** Saji Saraswathy Gopalan, Devi Kalyan Mishra, Ashis Kumar Das

**Affiliations:** Human Development Department, The World Bank, Washington DC, USA; Department of Community Medicine, Hitech Medical College, Rourkela, India; Research Group, The World Bank, Washington DC, USA

## Abstract

**Background:** Influenza could be associated with illnesses, severe complications, hospitalizations, and deaths among adults with high-risk medical conditions. Influenza vaccination reduces the risks and complications associated with influenza infection in high-risk conditions. We assessed the prevalence and predictors of influenza vaccination in a national sample of adults with high-risk medical conditions.

**Methods:** Using the nationally representative National Health Interview Survey of 2019, we estimated the prevalence of influenza vaccination among adults with high-risk conditions. We tested the associations between receipt of vaccination and sociodemographic predictors with adjusted multivariable logistic regression.

**Results:** Out of 15,258 adults with high-risk conditions, 56% reported receiving an influenza vaccine over the previous 12 months. Multivariable regressions show that respondents from older age groups, females, other race/ethnic group, married, higher annual family income, having a health insurance and those with more than two high-risk conditions are more likely to receive the influenza vaccine. However, adults from non-Hispanic Black race/ethnicity and living in the Southern census region are less likely to receive the vaccination. Education levels and living in a metro show no associations with vaccination status.

**Conclusion:** State authorities, primary physicians, specialists, and pharmacists have important roles in sensitizing and reminding individuals with high-risk conditions to receive timely vaccination. Similarly, affordability needs to be enhanced for influenza vaccination including better insurance coverage and reduced co-payment.

## Introduction

Influenza remains a serious infectious disease threat globally and in the United States.^1^ Several studies have shown that influenza could be associated with illnesses, severe complications, hospitalizations, and deaths among adults with certain chronic medical conditions.^2–4^ Additionally, it contributes to severe economic impact due to hospitalization and loss of workdays.^5^ Vaccination provides protection from influenza infection and related complications.^1^ For instance, vaccination averted an estimated 7.1 million illnesses, 3.7 million medical visits, 109,000 hospitalizations and 8,000 deaths among all age groups during the 2017-2018 flu season.^6^

Since 2010, the Center for Disease Control (CDC) and CDC’s Advisory Committee on Immunization Practices (ACIP) have been recommending routine annual influenza vaccination for all persons above six months old, who do not have any contraindications.^7^ Although routine vaccination is recommended for all ages, the existing evidence is limited on its prevalence among adults with high-risk health conditions. There are many studies on pregnant women and young children, but only a few on high-risk adult populations.^2,8–11^ The existing studies addressing vaccination prevalence among high-risk populations have mostly focused on a limited set of conditions and there is a need to explore others. Additionally, not many studies explored this among adults with multiple health risks. As comorbidity is a serious risk-factor, it would be relevant to understand the prevalence of influenza vaccination among adults with a number of high-risk medical conditions to inform relevant public health strategies. In this context, we assessed the prevalence and predictors of influenza vaccination among adults with high-risk medical conditions using a nationally representative National Health Interview Survey of 2019.

## Methods

### Data source

We analyzed data from the 2019 National Health Interview Survey (NHIS).^12^ The NHIS is a nationally representative annual household survey of the civilian noninstitutionalized population in the United States. It uses a multistage stratified cluster probability sampling design to obtain a nationally representative sample. In 2019, the NHIS content and structure were updated to better meet the needs of data users, improve the measurement of covered health topics, reduce respondent burden by shortening the length of the questionnaire, harmonize content with other federal health surveys, and incorporate advances in survey methodology and measurement.^12^ There were 31,997 sample adults and 9,193 sample children from 33,138 households in the 2019 data. The final sample adult response rate was 59.1%.^12^ We restricted our sample to adults aged 18 years and above with high-risk conditions. The high-risk conditions as reported by the respondents were: ever being told by a physician that they had coronary heart disease, angina, heart attack, stroke, diabetes, COPD, emphysema, chronic bronchitis, lymphoma, leukemia, or blood cancer; being diagnosed with cancer in the past 12 months (excluding non-melanoma skin cancer); hypertension in the past 12 months or under anti-hypertensive medications; reporting an asthma episode in the past 12 months; and extremely obesity (body mass index ≥40).

### Variables

The outcome of interest was self-reported receipt of an influenza vaccine during the previous 12 months prior to survey completion. The covariates were age in groups (18-39, 40-64, 65 and above), sex, race/ethnicity (non-Hispanic White, non-Hispanic Black, Hispanic, and others) education (below high school, high school, some college or higher), marital status (married and others), annual family income (<$35,000, $35,000-$74,999 and $75,000 and more), health insurance (none, public, private, and others), number of high-risk conditions (1, 2 and more than 2), residence (metropolitan and non-metropolitan), and the US census region (Northeast, Midwest, South, West).

### Statistical methods

We undertook descriptive analyses for the respondent characteristics and present the results stratified by subgroups for each covariate. The correlation was tested among all respondent characteristics with the Pearson’s correlation coefficient. Associations between receipt of influenza vaccine and predictors (age, sex, race/ethnicity, education, marital status, annual family income, health insurance, number of high-risk conditions, residence, and census regions) were estimated using a multivariable logistic regression model adjusting for all predictors. We considered the associations statistically significant if the p-value was below 0.05. Sampling weights were used to account for the multilevel design. The statistical analyses were performed using Stata Version 15 (StataCorp LLC. College Station, TX).

## Results

Table 1 shows the characteristics of individuals with high-risk conditions. There were 15,258 adults with high-risk conditions. Out of all, 43.5% were between 40 and 64 years, 53.4% were females and 47.9% were married. Sixty-five percent were of the non-Hispanic White race and 83.1% lived in metropolitan locations. Slightly over half had some college or higher education (55.1%) and had private health insurance (54.7%). There were 9% of respondents without any health insurance. Most (58.8%) had only one high-risk condition followed by 24.9% with two and 16.3% with more than two high-risk conditions. Around two-thirds (65.7%) were with cardio-vascular conditions. Among all adults with high-risk conditions, 56% reported receiving an influenza vaccine over the previous 12 months. Tables 2 shows that respondents with cancers and diabetes had the highest vaccination rates (65%) whereas those with extreme obesity had the lowest (46.9%).

**Table 1.** Sample characteristics of influenza vaccination among adults with high-risk conditions (N=15,258)

| **Characteristics** |  | **Unweighted**<br>**number** | **Weighted**<br>**proportion**<br>**(%)** |
| --- | --- | --- | --- |
| Age |  |  |  |
|  | 18-39 years | 2,350 | 21.3 |
|  | 40-64 years | 6,096 | 43.5 |
|  | >=65 years | 6,812 | 35.2 |
| Sex |  |  |  |
|  | Male | 6,866 | 46.6 |
|  | Female | 8,392 | 53.4 |
| Race/ethnicity |  |  |  |
|  | Non-Hispanic White | 10,735 | 65.0 |
|  | Non-Hispanic Black | 1,951 | 13.3 |
|  | Hispanic | 1,612 | 14.2 |
|  | Others | 960 | 7.5 |
| Education |  |  |  |
|  | Below high school | 1,848 | 16.0 |
|  | High school | 4,277 | 28.9 |
|  | Some college or higher | 9,133 | 55.1 |
| Marital status |  |  |  |
|  | Married | 8,414 | 47.9 |
|  | Not married | 6,844 | 52.1 |
| Annual family income |  |  |  |
| | <\$35,000 | 5,585 | 31.3 |
| | \$35,000-\$74,999 | 4,826 | 32.6 |
| | >=\$75,000 | 4,847 | 36.1 |
| Health insurance |  |  |  |
|  | None | 1,001 | 9.0 |
|  | Private | 8,221 | 54.7 |
|  | Public | 4,795 | 29.0 |
|  | Others | 1,241 | 7.3 |
| Number of high-risk conditions |  |  |  |
|  | 1 | 8,525 | 58.8 |
|  | 2 | 3,938 | 24.9 |
|  | >2 | 2,795 | 16.3 |
| Metro |  |  |  |
|  | Not metropolitan | 2,826 | 16.9 |
|  | Metropolitan | 12,432 | 83.1 |
| Census region |  |  |  |
|  | Northeast | 2,608 | 17.7 |
|  | Midwest | 3,379 | 20.8 |
|  | South | 5,884 | 40.0 |
|  | West | 3,387 | 21.5 |
| High-risk conditions*# |  |  |  |
|  | Extreme obesity | 3,765 | 27.7 |
|  | Cancers | 674 | 3.9 |
|  | Diabetes | 3,292 | 20.8 |
|  | Cardio-vascular diseases | 10,756 | 65.7 |
|  | Lung diseases | 3,738 | 24.8 |
| Received influenza vaccine |  |  |  |
|  | No | 6,089 | 44.0 |
|  | Yes | 9,169 | 56.0 |

\* Extremely obesity – body mass index $\geq 40$ ; cancers include solid and hematological tumors; cardio-vascular diseases include coronary heart disease, angina, heart attack, stroke, hypertension; lung diseases include COPD, emphysema, chronic bronchitis, asthma

**Table 2.** Multivariable logistic regression of influenza vaccination among adults with high-risk conditions by socio-demographic characteristics (N=15,258)

| Characteristics |  | Proportion<br>received<br>vaccine<br>(%) | Adjusted<br>odds<br>ratio | 95%<br>confidence<br>interval | p value |
| --- | --- | --- | --- | --- | --- |
| Age |  |  |  |  |  |
|  | 18-39 years | 38.5 | Reference |  |  |
|  | 40-64 years | 50.6 | 1.20 | 1.05-1.37 | 0.006 |
|  | >=65 years | 73.2 | 2.96 | 2.56-3.43 | <0.001 |
| Sex |  |  |  |  |  |
|  | Male | 53.8 | Reference |  |  |
|  | Female | 57.8 | 1.30 | 1.19-1.42 | <0.001 |
| Race/ethnicity |  |  |  |  |  |
|  | Non-Hispanic White | 59.1 | Reference |  |  |
|  | Non-Hispanic Black | 46.4 | 0.74 | 0.64-0.84 | <0.001 |
|  | Hispanic | 48.1 | 0.96 | 0.83-1.12 | 0.641 |
|  | Others | 60.2 | 1.24 | 1.03-1.50 | 0.022 |
| Education |  |  |  |  |  |
|  | Below high school | 53.7 | Reference |  |  |
|  | High school | 51.7 | 0.92 | 0.79-1.08 | 0.296 |
|  | Some college or higher | 58.8 | 1.16 | 0.99-1.35 | 0.061 |
| Marital status |  |  |  |  |  |
|  | Not married | 51.8 | Reference |  |  |
|  | Married | 59.7 | 1.13 | 1.03-1.24 | 0.013 |
| Annual family income |  |  |  |  |  |
| | <\$35,000 | 52.6 | Reference | | |
| | \$35,000-\$74,999 | 54.9 | 1.08 | 0.96-1.21 | 0.19 |
| | >=\$75,000 | 59.8 | 1.35 | 1.18-1.55 | <0.001 |
| Health insurance |  |  |  |  |  |
|  | None | 27.6 | Reference |  |  |
|  | Private | 56.8 | 2.22 | 1.83-2.71 | <0.001 |
|  | Public | 61.0 | 2.10 | 1.70-2.58 | <0.001 |
|  | Others | 64.7 | 2.76 | 2.16-3.51 | <0.001 |
| Number of high-risk conditions |  |  |  |  |  |
|  | 1 | 51.4 | Reference |  |  |
|  | 2 | 59.4 | 1.07 | 0.93-1.23 | 0.331 |
|  | >2 | 67.2 | 1.22 | 1.00-1.49 | 0.046 |
| Metro |  |  |  |  |  |
|  | 0 | 55.6 | Reference |  |  |
|  | 1 | 56.0 | 1.04 | 0.93-1.17 | 0.441 |
| Census region |  |  |  |  |  |
|  | Northeast | 60.0 | Reference |  |  |
|  | Midwest | 57.2 | 0.93 | 0.80-1.06 | 0.277 |
|  | South | 52.2 | 0.82 | 0.72-0.93 | 0.002 |
|  | West | 58.4 | 0.97 | 0.84-1.12 | 0.659 |
| High-risk conditions*<sup>#</sup> |  |  |  |  |  |
|  | Extreme obesity | 46.9 | 1.11 | 0.96-1.28 | 0.167 |
|  | Cancers | 65.1 | 1.26 | 1.08-1.47 | 0.004 |
|  | Diabetes | 65.0 | 1.36 | 1.19-1.57 | <0.001 |
|  | Cardio-vascular diseases | 61.4 | 1.19 | 0.94-1.51 | 0.148 |
|  | Lung diseases | 55.4 | 0.87 | 0.76-1.00 | 0.048 |

\* Extremely obesity – body mass index $\geq 40$ ; cancers include solid and hematological tumors; cardio-vascular diseases include coronary heart disease, angina, heart attack, stroke, hypertension; lung diseases include COPD, emphysema, chronic bronchitis, asthma
<sup>#</sup> Reference category for the high-risk conditions are those without the condition as some respondents have multiple conditions

There were not strong correlations between predictors with the Pearson’s correlation coefficients ranging from −0.21 to 0.39. Multivariable regressions show that (table 2) respondents from older age groups, females, other race/ethnic group, married, higher annual family income, having health insurance, and those with more than two high-risk conditions are more likely to receive the influenza vaccine. In addition, those with cancers and diabetes have a higher likelihood of being vaccinated. However, adults from non-Hispanic Black race/ethnicity, living in the Southern census region, and having lung diseases are less likely to receive the vaccination. Education levels and living in a metro were not associated with the receipt of the vaccine.

## Discussion

We assessed influenza vaccination rates among various sociodemographic groups in a nationally representative sample of US adults with high-risk conditions. Only 56% of adults with a high-risk condition received influenza vaccination in the prior 12 months falling short of the Healthy People 2020 target of 70% set for all adults. Data from National Immunization Survey-Flu and Behavioral Risk Factor Surveillance System (BRFSS) from 2010-11 through 2018-19 influenza seasons show that the vaccination rates among adults (18-64 years) with high-risk conditions have historically remained lower than 50% (range 38.8%-47.9%).^13^ The higher vaccination rate in our study could be due to two reasons. First, we used a different data source –the NHIS data. During the 2018-19 flu season, the estimated influenza vaccine coverage rates among adults were 5% points higher for the NHIS data compared to BRFSS data.^14^ Second, we included persons above 64 years in our study. Our analysis shows older age groups have a higher likelihood of receiving the vaccine. A few other studies have explored influenza vaccination among adults with high-risk conditions from multiple data sources. Using the NHIS, a study has shown the coverage at 49.5% among 18-64 years for the 2012-13 season.^15^ Another study using the BRFSS survey of 2013 found the vaccination rates between 47.4% and 57.8% for adults with high-risk conditions in Kansas state.^10^ The annual vaccination rate was 47.3% in a nationally representative sample of non-Hispanic Black and White high-risk adults in an online survey in 2015.^16^

Our study found a higher rate than what CDC has reported using BRFSS (45.3%) for the overall adult population above 18 years for the season 2018-2019.^14^ However, these estimates indicate a slight increment in the annual overall vaccination rate among adults over the last few years. It was 45.3% in 2018-2019, an increase of 8.2% points from the 2017-2018 flu season and 2% points higher than 2016-2017. Perhaps the high-risk groups could have followed the general trends in the vaccination receipt for the year 2018-2019. Although further improvements are needed, the recent years have witnessed intense efforts from the disease-related programs to encourage vaccination receipt to improve the health of people with chronic conditions.^17^ These efforts could have contributed to the increased vaccination rate among high-risk groups.

Vaccination rates in our study were significantly lower among men, younger adults (18-39 years), non-Hispanic Blacks, individuals without insurance, and those from low-income families. These disparities are similar to those reported in earlier studies of adults with high-risk conditions.^9,11,15,16^ A 2017 study found that psychosocial factors play a major role in vaccination receipt and the non-Hispanic Black population reported a low self-perceived social status and had low vaccination.^16^ It additionally reports that social groups with a historically low prevalence of vaccinations tend to have a lower chance to be vaccinated. This requires culturally appropriate and targeted health awareness strategies.

We found people with more than two high-risk conditions had a higher chance for vaccination compared to those with lower number of high-risk conditions. A 2013 study among 18-64 years old high-risk groups found similar trends, it reported 49.5% vaccination rate among those with one health risk and 59.5% among people with more than one risk.^15^ As mentioned earlier, the encouragement from the disease-related programs could be a positive contributor here. In accordance with the current evidence, our study finds similar vaccination rates for adults with diabetes, but lower for cardio-vascular diseases.^9,18^

We found a lower vaccination chance among people with no insurance. This finding calls for increased affordability for influenza vaccination and reduced copayment, especially for low-income groups. People with Medicare can get the flu shot at no cost.^19^ However, the encouragement from the routine providers would be a decisive factor to avail flu shots among Medicare beneficiaries. A study found that 87.3% with high-risk conditions who did not vaccinate for the 2012-2013 season reported more than one hospital visit.^15^ We could not verify in our data how was the vaccination receipt in counties that provide free vaccination. This could have possibly explored to what extent affordability and consumer choices interact on vaccination behavior. However, we presume that free availability alone is not enough rather adequate awareness regarding the free availability and health risks of influenza would be essential. A recent study found that the perception of a higher risk of contracting influenza among those with chronic conditions is a predictor for vaccination.^16^

As high-risk populations are on routine care, constant and frequent encouragement from their providers in the influenza season would increase the uptake. This requires a further stronger engagement of the state vaccination programs with the providers and practices. A study in 2013 found that a substantial portion of adults who made 4-10 visits were not vaccinated.^15^ One study reported that 22% of subspecialists did not stock the influenza vaccine and recommend one.^20^ Intense encouragement would be needed from primary physicians, specialists, and pharmacists. They could also take proactive actions in referring patients for vaccination.

Findings also call for further strengthening of the ongoing efforts of the disease-related programs and the Department of Health and Human Services on encouraging vaccination among high-risk groups. Intensifying the ongoing health education strategies and reminder notifications through multiple platforms such as television, radio, telephone, and social media could be beneficial.^21^

More than generic messages, detailing the increased risks of missing the vaccination among high-risk groups could be emphasized. The ongoing health awareness programs for chronic conditions (e.g., cancer, diabetes, asthma) could also strengthen the frequency and platforms of sensitization for influenza vaccine by including thematic awareness messages for specific risk groups.

The major limitation of this study is the dependence on self-reported information leading to potential recall bias. However, self-reported seasonal influenza vaccination has shown a high agreement with the data from medical records.^22,23^ Despite this, we cannot ignore the possibility of either over- or under-estimation of the true prevalence of the vaccination. Due to the larger sample sizes, data quality and representativeness of the NHIS data, the study findings will be relevant to strengthen the ongoing health awareness strategies and affordability for the influenza vaccine to improve the health of high-risk groups.

## Conclusions

Influenza vaccination coverage among the high-risk adult population was 56% in 2019. Younger adults (<40 years), males, non-Hispanic Black race/ethnicity, lower annual family income, without health insurance, and those with less than two high-risk conditions are less likely to receive the influenza vaccine. State authorities, primary physicians, specialists and pharmacists have important roles in sensitizing and reminding individuals with high-risk conditions to receive timely vaccination.

## Conflicts of interest

The authors declare that there is no conflict of interest. The views expressed in the paper are that of the authors and do not reflect that of their affiliations.

## Funding statement

This study did not receive funding from any source.

## Data Availability

Data from publicly available NHIS study

## Acknowledgements

We are grateful to the National Health Interview Survey for making this data publicly available.

## Notes

### Competing Interest Statement

The authors have declared no competing interest.

### Funding Statement

No external funding was received

